# A single-cell atlas depicting the cellular and molecular features in human ligamental degeneration: a single cell combined spatial transcriptomics study

**DOI:** 10.1101/2023.01.05.23284219

**Authors:** Runze Yang, Tianhao Xu, Lei Zhang, Minghao Ge, Liwei Yan, Jian Li, Weili Fu

## Abstract

**Background:** To systematically identify cell types in human ligament, investigate how ligamental cell identities, functions, and interactions participated the process of ligamental degeneration and explore the changes of ligamental microenvironment homeostasis in the disease progression.

**Methods:** Using single-cell RNA sequencing and spatial RNA sequencing of approximately 49356 cells, we created a comprehensive cell atlas of healthy and degenerated human anterior cruciate ligaments. We explored the variations of the cell subtypes’ spatial distributions and the different processes involved in the disease progression, linked them with ligamental degeneration process using computational analysis.

**Results:** We identified new fibroblast subgroups contributed to the disease and mapped out their spatial distribution in the tissue and revealed two stages of the degenerative process. We compared the cellular interactions between different tissue states and identified important signaling pathways may contribute to the disease.

**Conclusion:** This cell atlas provides the molecular foundation for investigating how ligamental cell identities, biochemical functions, and interactions contributed to the ligamental degeneration process. The discoveries revealed the pathogenesis of ligamental degeneration at single-cell and spatial level which is characterized by extracellular matrix remodeling. Our results provide new insights into the control of ligamental degeneration and potential clues to developing novel diagnostic and therapeutic strategies.

**Funding:** This study was funded by the National Natural Science Foundation of China (81972123, 82172508), Sichuan Science and Technology Program (2020YFH0075), Fundamental Research Funds for the Central Universities (2015SCU04A40), Chengdu Science and Technology Bureau Project (2019-YF05-00090-SN), and 1.3.5 Project for Disciplines of Excellence of West China Hospital Sichuan University (ZYJC21030, ZY2017301).

## Introduction

With the increasing aging of the world’s population, most of us crave a healthy and long life(Beard & Bloom, 2015). Good musculoskeletal healthy is essential to people to live independently in the society throughout their life course(Briggs et al., 2016). Skeletal ligaments, an important part of the musculoskeletal, are defined as dense connective tissue that span the joint and then fixed at both ends of the bone. One of the main functions of ligaments is mechanical as they passively maintain joint stability and assist in guiding of those joints through their normal range of motion when a tensile loading is applied(Frank, 2004). The knee is the largest and most complicated hinge joint associated with weight bearing in the human body. The anterior cruciate ligament (ACL) is crucial for knee kinematics, especially in rotation, and functions as an anterior/posterior stabilizer(Fleming, 2003). ACL degeneration can gradually result in chronic knee pain, instability and even poor life quality(Thompson et al., 2015). ACL rupture is a risk factor for cartilage degeneration and the development of osteoarthritis (OA)(Roos, Adalberth, Dahlberg, & Lohmander, 1995). It has been reported that aging-related degenerative changes in the ACL might also contribute to OA occurrence and progression(Loeser, 2010), but mechanisms of ACL aging and degradation are remain unclear.

The extracellular matrix (ECM) is the physical basis for the biological functions of ligaments, and ACL degeneration is accompanied by alterations in the ECM. The ECM of ACL consist of collagen types I, II, III, and V, elastin, and proteoglycans(Laurencin & Freeman, 2005), and collagen type I is a major determinant of tensile strength(Corps et al., 2006). ACL degeneration is characterized by disorganization of collagen fibers, cystic changes, mucoid degeneration and chondroid metaplasia(Hasegawa et al., 2012). Different types of pathological changes correspond to different ECM alterations. For example, mucoid degeneration reflects degradation of collagen and deposition of new glycosaminoglycans and cystic changes represent devoid of ECM in the diseased area(Hasegawa et al., 2012). It has been reported that some regions in the degenerated ACL have decreased type I collagen, whereas type II, III, and X collagen and aggrecan are significantly increased, and this abnormal ECM production can lead to biomechanical fault(Hasegawa et al., 2013; Hayashi et al., 2003). In recent years, many scholars have paid attention to the degeneration mechanism of ligaments. TGFβ1, a member of the TGF superfamily, can be used to induce chondrogenic differentiation of ligament-derived stem cells in vitro(Schwarz et al., 2019) and is considered to be an essential molecule contributing to chondroid metaplasia. Some reports suggested that complement cascade could lead to ECM catabolism and contribute to ligament degeneration(Busch et al., 2013; Schulze-Tanzil, 2019).

There are various types of cells in the ligament(Kharaz, Canty-Laird, Tew, & Comerford, 2018) and building a detailed ligamental cell landscape is essential to understand ligamental characteristics and underlying pathogenesis of ACL degeneration and OA. As a result of the continuous technological innovation, single-cell RNA sequencing (scRNA-seq) is recognized as a significant tool for depicting cellular heterogeneity(Wen & Tang, 2018; Zeng et al., 2019). Spatial RNA sequencing (spRNA-seq) is a recently developed revolutionary technology. It combined the advantages of the comprehensive analysis of bulk RNA sequencing and in situ hybridization to provide complete transcriptome data with spatial information(Li & Wang, 2021). Here, we performed scRNA-seq and spRNA-seq to obtain an unbiased atlas of ACL cell clusters. Our findings provide a better understanding of the inherent heterogeneity, construct the classifications of fibroblasts and profile the spatial information of identified cell clusters in the ACL. Notably, we also identified the cell interactions during the disease process and demonstrate the role of FGF and TGF-β signaling pathways in ligament degeneration. Thus, our study reveals the cellular landscape of the human ACL and provides insight that could help to identify therapeutic targets for human ligamental degeneration.

## Methods

### Human ligament cell sample preparation

The ligament specimens were collected from four joint replacement patients with osteoarthritis (degenerated ACLs) and four amputation patients with osteosarcoma (healthy ACLs).

All specimens removed were placed immediately in Dulbecco modified Eagle medium (DMEM) free of antibiotics and FBS under 4°C. Ligament samples were rinsed in precooled PBS and then were cut into 1 mm^3^ pieces. Containing Collagenase type I (2 mg/ml) and 0.25% (w/v) trypsin DMEM was used to digest the specimens for 1 h at 37°C using a Thermomixer at 1200 rpm (Eppendorf, Hamburg, Germany). After that, Ham’s F-12 media containing 10% FBS was added to stop the process of digestion and then the digested tissue passed through a 100 μm cell strainer. Finally, after centrifugation, cells were collected for subsequent batch analysis.

### Single-cell RNA-seq analysis

The raw single cell sequencing data was mapped and quantified with the 10 × Genomics Inc. software package CellRanger (v5.0.1) and the GRCh38 reference genome. Using the table of unique molecular identifiers produced by Cell Ranger, we identified droplets that contained cells using the call of functional droplets generated by cell ranger. After cell containing droplets were identified, gene expression matrices were first filtered to remove cells with > 5% mitochondrial genes, < 250 or > 8000 genes, and < 500 UMI. Downstream analysis of Cellranger matrices was carried out using R (4.1.3) and the Seurat package (v 4.1.0, satijalab.org/seurat). In total, 49356 cells with an average of 2435 genes per cell were selected for ongoing analysis. Of these single cells,24721 were obtained from lesions ligament, included L2, L5, L6 and L8. The remainder were cells from healthy and included L1, L3, L4 and L10.

### Single-cell trajectory analysis

We used the Monocle3 v. 2.8.0 R package to infer a hierarchical organization part of fibroblasts, to organize these cells in pseudotime. We took these subpopulations from the Seurat data set from which we reperformed shared nearest neighbor clustering and differential expression analysis as described previously. We then selected the differentially expressed genes based on fold-change expression for Monocle to order the cells using the DDRTree method and reverse graph embedding. We find gene co-expression modules according to the trend of gene expression, and show the relationship between cells and modules in the form of heat maps.

### Ligand-receptor interaction model

We used the ligand-receptor interaction database from cellchat database to determine potential ligand-receptor interactions between fibroblast subpopulations and other cell types. The expression data were preprocessed for subsequent cell-cell communication analysis. The ligand or receptor that is overexpressed in a class of cells is first identified, and the gene expression data is then projected into a protein-protein interaction network. Whenever a ligand or receptor is overexpressed, the ligand-receptor interaction is recognized. CellphoneDB is based on the expression of a receptor in one cell type with a ligand in another cell type, resulting in rich receptor-ligand interactions between two cell populations. For the gene expressed by the cell population, the percentage of cells expressing the gene and the average gene expression were calculated. The gene was removed if it was expressed only in 10% or less of the cells in the population (the default value is 0.1).

### Spatial-transcriptome analysis

We used L1, L8 samples for spatial transcriptome analysis. We performed the Seurat standard analysis. Machine prediction and marker gene methods were used to determine the cell type and used SCTransform for standard analysis of the data. First, we determine it is specific enough to finding a topic profile (similar to a feature vector) for each cell type, and then deconvolved the information by cell type and superimposed it onto the slice in pie chart form.

### Deconvolution analysis

SCDC (v 0.0.0.9000) tool combined with single cell data was used to deconvolve bulk RNAseq data for cell type and content proportion analysis bulk RNAseq data. The bulk RNAseq data is also our own data. Histogram and heat map can be drawn for the predicted data, and the proportion of cell type content can be compared between groups. Then we used GSVA (v 1.42.0) to calculate the same data and draw heat maps to aid the analysis.

### Statistical analysis

Nonparametric Wilcoxon rank sum test was used to analyze the differences between two groups. All statistical analyses were performed in R or GraphPad Prism (version 5.0). Statistical significance was defined as *p < 0.05, **p < 0.01, ***p < 0.001.

## Results

### 1. Comprehensive scRNA-seq analyses resolve the major cell types in the human normal and degenerated ligament

To decipher cellular heterogeneity and construct the cell landscape for ligament degeneration, we performed single-cell transcriptomic profiling of cells from four healthy and four degenerated ACL. The characteristic pathological findings in the ACL of OA were identified by arthroscopy (figure1 A and B). In the normal group, the ligaments are white stripes with clear structure, and blood vessels can be seen on their surfaces. In the degenerated group, the ligaments were swollen, the structure was disorganized, and the color of the tissue was dark. From the images of knee MRI, we can also find that the signal of normal ACL on T2 phase is uniform and the structure is clear, while the signal of the ACL in the degenerated group is mixed, and the clear structure cannot be distinguished (supplementary figure 1 A and B). After rigorous quality control, we obtained the transcriptomes of 49356 cells (normal: 24635 degenerated: 24721) and then employed differential gene expression analysis to discern cluster-specific markers. We firstly used principal component analysis (PCA) to reduce the dimension, and then we adopted uniform manifold approximation and projection (UMAP) method to conduct next analysis. Unbiased clustering based on UMAP identified four cell classes and histrionic lineage-defining genes were detected, including endothelial cells: PECAM1, VWF, and PLVAP (3862, 7.83%), fibroblasts: DCN, LUM, and COL1A2 (32644, 66.14%), immune cells: PTPRC, SRGN, and CD163 (8021, 16.25%), and pericytes: ACTA2, MCAM, and RGS5 (4829, 9.78%) (figure 1 C-F and supplementary figure1 C). According to figure1 I, we were able to further confirm that the selected cell markers could well identify various cell types. UMAP plots and stacked bar plots in figure 1G described the distributions of cells in the healthy and degenerated samples. We can conclude that, in general, fibroblasts are the main cell type in both normal and degenerated ligaments and the degenerated group had higher immune cells ratio and lower pericytes ratio compare with the normal group. We next analyzed the number of differentially expressed genes (Degs) between healthy and degenerated ligament clusters. The results demonstrated that the fibroblast had the largest difference, implying that fibroblasts undergo significant changes during the degenerative progress (figure 1 H). We also collected normal as well as degenerated ligament specimens for bulk sequencing and performed deconvolution analysis between bulk sequencing results and scRNA-seq results. The results illustrated that scRNA-seq samples were in good agreement with bulk sequencing samples (figure 1 J and K). After that, we verified the top 5 genes that were highly expressed in the degenerative state of the 4 cell subsets in the bulk sequencing results and these results also have a high consistency (supplementary 1 D-G).

**Figure 1:**
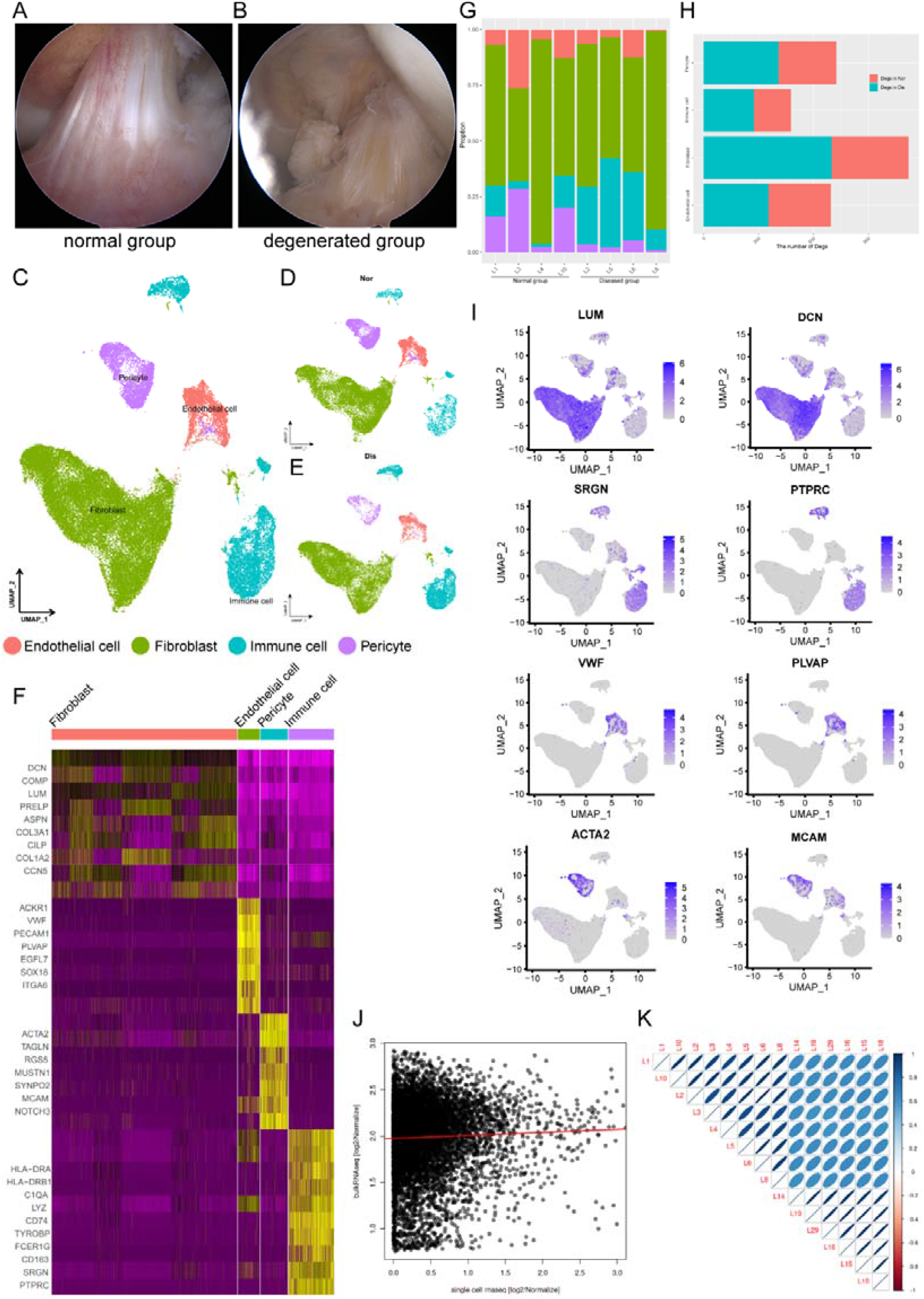
Single-cell RNA-seq reveals major cell classes in human ligament A and B: A photograph of typical normal (left) and degenerated (right) anterior cruciate ligaments (ACL) under arthroscope. C: UMAP visualization of all cell clusters in collected ligamental specimens. D and E: UMAP visualization of the donor origins in normal/diseased samples. F: Heatmap of selected marker genes in each cell cluster. G: The percentages of the identified cell classes in normal/diseased ligament. H: Number of differentially expressed genes (DEGs) in each cell type of normal/diseased status. I: Feature plots of expression distribution for selected cluster-specific genes. Brighter colors indicate higher expression levels. J: Gene expression profiles from bulk samples (n = 6) and in single cell samples (n = 8) were averaged and plotted on X and Y axes, respectively. Red lines indicate linear model fit and the diagonal. K: Correlation heatmap shows Pearson’s correlation between all bulk and in single cell samples.

### 2. Characterizations of subpopulations of ligamental fibroblasts between different kinds of tissue states

Because fibroblasts are known to play a vital role in ECM homeostasis and the degeneration of the ligament and undergo remarkable changes during the process of ligament degeneration, we next conducted further analysis of fibroblasts in normal and degenerated ligaments. We subclustered fibroblasts and identified 10 subsets (Fib.1-Fib.10) (figure 2A). The distribution shown in figure 2B indicated that the batch effects are resolved well. As illustrated in figure 2C, we obtained the cell proportions of the fibroblast subpopulations between normal and diseased groups. We can conclude that the ratio of fibroblast subclusters varied greatly between these two groups. The ratio of Fib.1, Fib.2, Fib.8, and Fib.9 in the diseased group was obviously higher than that in the healthy group, and the ratio of Fib.3, Fib.4, and Fib.7 in the healthy group was obviously higher compare to the diseased group. The proportion of remaining fibroblast subclusters did not differ significantly between the two groups.

**Figure 2:**
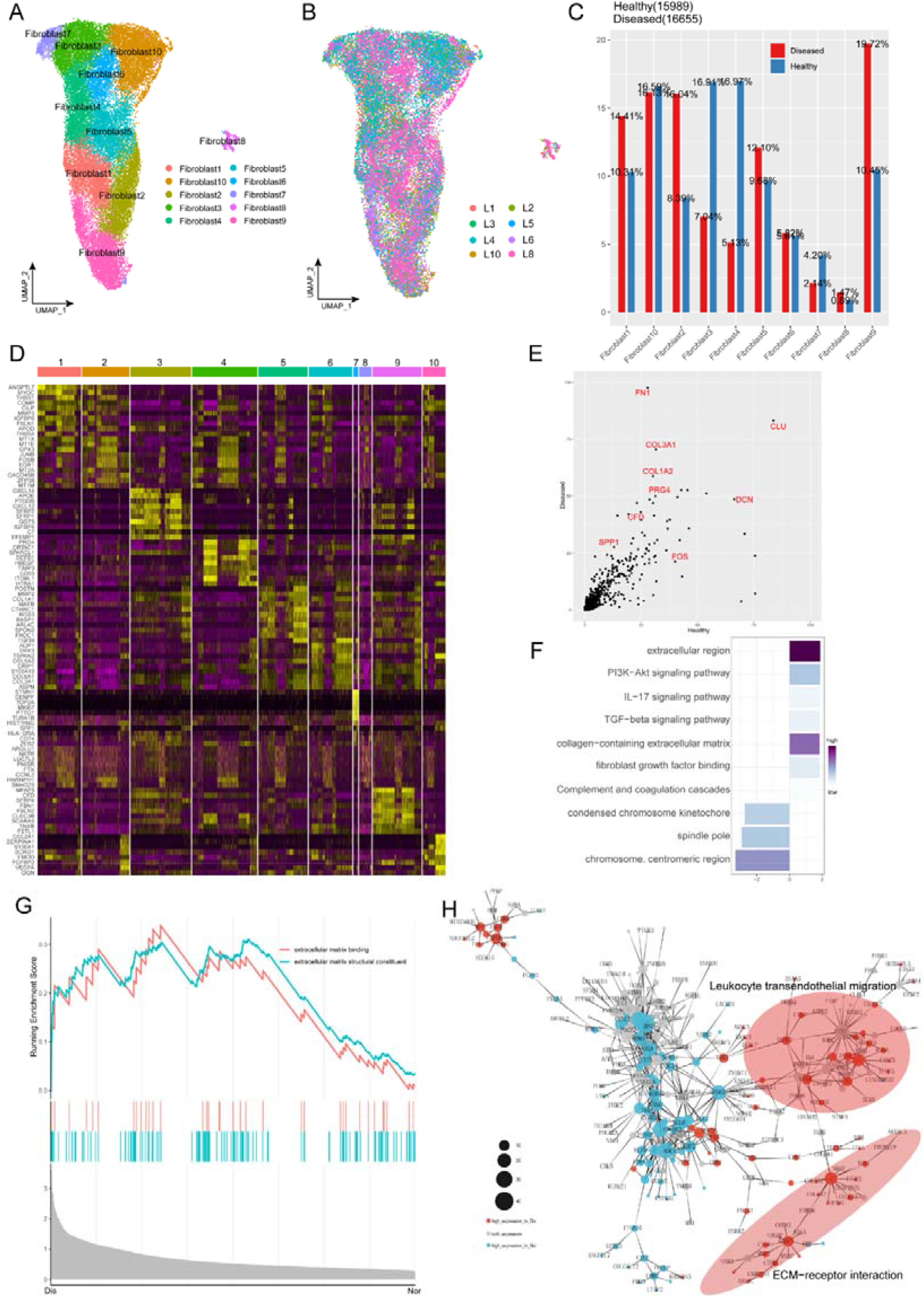
Characterization of fibroblast subclusters across different statuses in human ACL A: UMAP visualization of the subclusters of fibroblasts. B: UMAP visualization of the distribution of fibroblast subclusters at different samples. C: The proportions of 10 fibroblast subpopulations in normal and diseased ligaments. D: A cell-level heatmap reveals the normalized expression of DEGs for each fibroblast cluster defined in. E: Volcano plot showing the DEGs between two statues of ligamental fibroblasts. The x axis represents highly expressed genes in normal cells, and the y axis represents highly expressed genes in diseased cells. F: GO and KEGG enrichment analysis of DEGs in ligamental fibroblasts between normal and degenerated states. G: GSEA enrichment plots for representative signaling pathways upregulated in fibroblasts of diseased samples, compared with normal samples. H: Gene-gene interaction networks between DEGs in ligamental fibroblasts of normal group and fibroblasts of diseased group.

According to the analysis of Degs among these fibroblast subgroups, we tried to reveal the identity of each subpopulation (figure 2D). Combined with the analysis of the proportion of cell subclusters, we inferred that Fib.1 and Fib.2 are ECM remodeling associated fibroblasts as they highly expressed the genes related to ECM components and ECM decomposition such as COL1A1, CTHRC1, COL3A1, FBLN1, MMP2, MMP14 and TPPP3. From the perspective of ECM synthesis, Fib.1 and Fib.2 are slightly different. Fib.1 highly expressed the collagen related genes, but Fib.2 highly expressed the genes related to synthesis of connexins. Fib.3 expressing metallothionein family genes such as MT1X, MT1E, and MT1M is a population of homeostasis-associated fibroblasts with defensive functions. Fib.4 expressing genes associated with formation and organization of the ligamental ECM such as ANGPTL7, MYOC, CILP2 and THBS1 is a group of resident fibroblasts that function normally. Fib.5 is a cluster of structural fibroblasts within the ligament as they highly expressed multiple collagen-related genes such as COL3A1, COL6A1, COL5A1, COL1A1, and TGFVBI. Fib.6 is a cluster of chondrocytes as they highly expressed cartilage related genes such as COL2A1, FMOD, CHAD, ACAN, and CILP2. We cannot define Fib.7 well, because they expressed Degs without specific tags. Fib.8 is a group of cycling cells as they highly expressed cell cycle related genes such as CENPF, TOP2A, and MLI67. Fib.9 is a group of inflammation related fibroblasts as they highly expressed CXCL14, CXCL12, C3, and C7. Fib.10 may have the function of damage repair as they highly expressed anti-inflammation and anti-ECM decomposing associated genes such as PRG4, DEFB1, TIMP3, and HBEGF.

For further in-depth investigation of changes in ligamental fibroblasts during the degeneration process, we performed the Degs analysis in the whole fibroblast population between the normal and degenerated ligaments. We observed genes upregulated in the degenerated group such as CFD, SPP1, COL1A2 and COL3A1, which were related to scar healing and ECM remolding (figure 2E). Gene Ontology (GO) and Kyoto Encyclopedia of Genes and Genomes (KEGG) analysis were conducted for further functional interpretation of these Degs. The results illustrated that the upregulated genes in diseased group were enriched for the terms “extracellular region”, “TGF-beta signaling pathway” and “Complement and coagulation cascades” (figure 2F). Gene Set Enrichment Analysis (GSEA) also demonstrated that ECM associated signaling, ECM binding and ECM structural constituent, were activated in the degenerated group, which implied that the changes of ECM play an important role in the ligament degeneration (figure 2G). And then by building gene interaction networks, we identified that genes upregulated in degenerated groups were concentrated in two parts, ECM-receptor interaction and leukocyte trans-endothelial migration, which once again proved the significant function of ECM remolding and immune inflammation in the disease process (figure 2H).

### 3. Dynamic transcriptional changes in ligamental degeneration

To gain insight into the cellular progression of fibroblast subclusters during the disease process, we performed cell trajectory analysis, revealing that two significant routes in the process of degeneration. Five fibroblast subclusters, including Fib.2, Fib.4, Fib.5, Fib.9, and Fib.10, were involved in reconstruction the disease trajectories using Monocle 3, an algorithm for the reconstruction of lineage programs based on similarity at the transcriptional level(Cao et al., 2019). We set Fib.4 and Fib.5 as the starting point of the trajectories due to they are structural fibroblasts with high expression of ECM component genes and represent normal ligamental functions well, and then computed pseudotime for cells along the inferred developmental axis (figure 3A and B). More specifically, Fib.4 and Fib.5 were predicted to transformed into two distinct cell fate, including the cell fate1, which includes Fib.2 and Fib.9, and the cell fate2, which includes Fib.10. With this in mind, we tried to explore the genic dynamics that distinguished these two cell fates. The expression profile of cell fate1 showed increasingly high expression of genes (CXCL12, MMP2, MMP14, C7) related to “leukocyte trans-endothelial migration”, “cellular response to chemokine”, and “inflammatory response pathway”. Along with the cell fate2, we observed gradually high expression of ECM organization and skeletal muscle tissue development related genes (FOS, EGR1, FGF10, FN1) (figure 3C-H).

**Figure 3:**
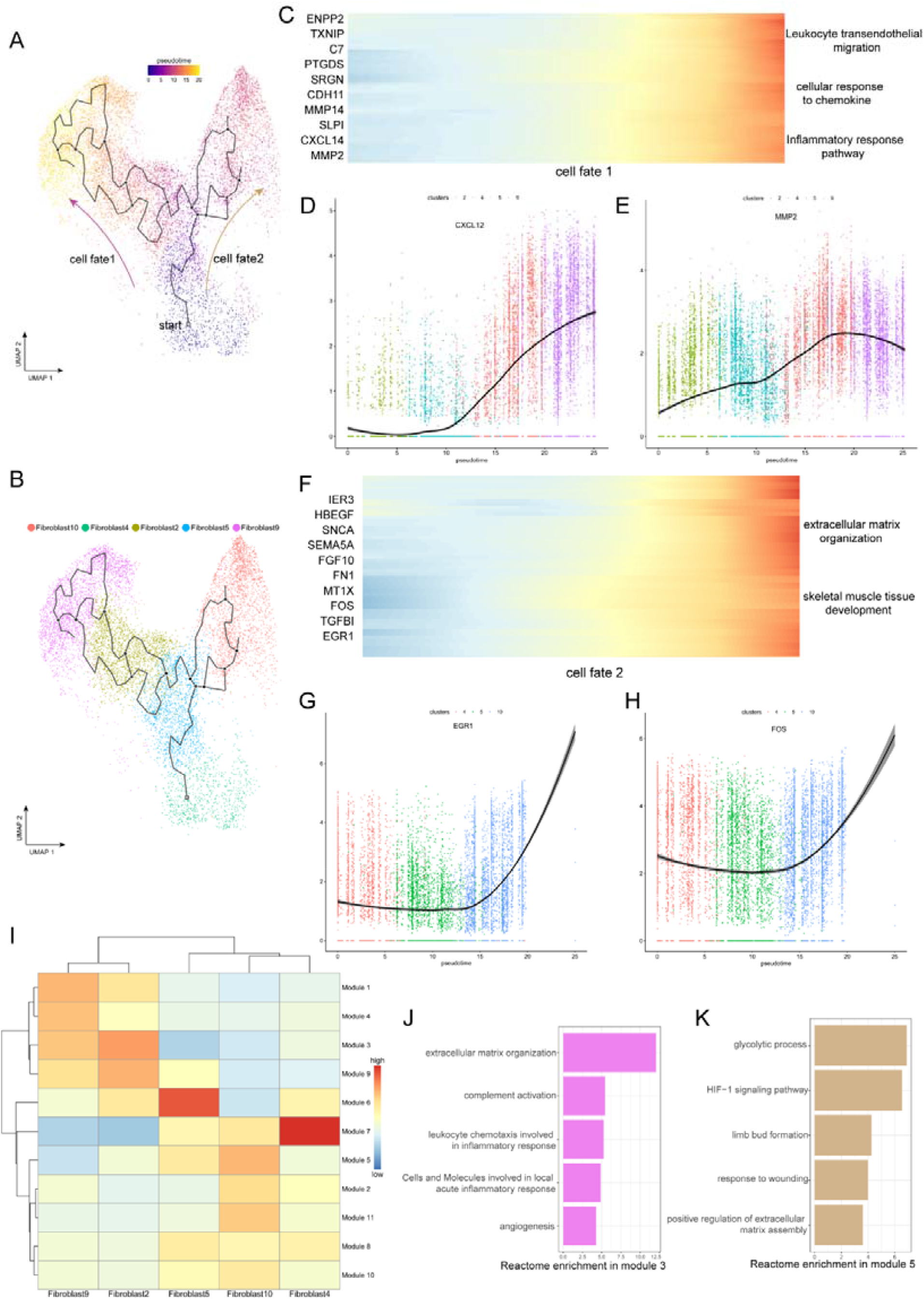
Evolution trajectory and transcriptional fluctuation during ligamental degeneration progression A and B: UMAP visualization of fibroblast 2,4,5,9,10. Developmental pseudotime for cells present along the trajectory inferred by Monocle 3, with cell fate1 and cell fate2 branches coming from fibroblast 4. C: Heatmap showing the expression changes of the highly variable genes along the cell fate1 in normal and degenerated groups. D and E: Representative gene expression levels along cell fate1 trajectory of normal and diseased statuses. F: Heatmap showing the expression changes of the highly variable genes along the cell fate2 in normal and degenerated groups. G and H: Representative gene expression levels along cell fate2 trajectory of normal and diseased statuses. I: Heatmap showing the scaled mean expression of modules of coregulated genes grouped by Louvain community analysis across the subclusters. J and K: Enrichment analysis results of Module 3 and 5.

To investigate gene expression dynamics along the trajectories, we group genes that varied between cell clusters into Module 10 using Louvain community analysis. We can observe the aggregated expression of each module in the figure 3I. We identified that the expression of genes in module 3 was gradually elevated along the cell fate1 and gradually declined along the cell fate2, which were enriched for genes related to rheumatoid arthritis, antigen processing and presentation and complement activation. In contrast, the expression of wound repair and ECM assembly related genes were gradually elevated along the cell fate2 but declined along the cell fate1, such as ADAM12, HS3ST3A1, FN1, and SERPINE2 in module 5 (figure 3I-K). According to these pseudotime analysis results, we inferred that there are two opposite dynamic trajectories in the process of ligamental degeneration, one is the progressive damage process of complement inflammation leading to the continuous progression of the disease, and the other is the process of repairing the damage to delay or even reverse the disease process.

### 4. Characterization of stromal cells in coordinating ligament microenvironment during degeneration progression

Besides fibroblasts, various endothelial cells (ECs) and immune cells also play significant roles in the occurrence and development of ligament degeneration. In our ligament samples, we identified seven clusters of blood-vessel derived cells including five clusters of ECs (EC1-5) and two clusters of pericytes (pericyte1 and pericyte2). Fig 4A shown the distribution of EC1 expressing high level of ACKR1 is a population of venous ECs; EC2 highly expressed COL4A1, COL4A2, H19 was related to anti-angiogenesis; EC3 highly expressed genes associated with ECM organization and we inferred this cluster related to damage repair. EC4 expressing high level of CCL2, CCL8, STEAP4 and SYNPO2 is a cluster of ECs related to inflammatory chemotaxis. EC5 is a cluster of lymphatic ECs. As for pericytes, we identified two populations expressed pericyte markers ACTA2, MCAM. Pericyte1 highly expressed myocyte related genes such as MYH11, MUSTN1 and LBH, which implied that this cluster may have myocyte-like properties. Pericyte2 highly expressed fibroblast markers such as COL1A1 and COL3A1, which implied that this cluster may have fibroblast-like properties (figure 4G). We next compared the proportions of these 7 clusters between normal and diseased ligament (figure 4Cand D). The results illustrated that most ECs subsets were elevated in the diseased group, implying increased angiogenesis during ligamental degeneration. The ratio of pericyte1 in the diseased group was higher than that in the healthy group, but the ratio of pericyte2 was lower. Increased ratio of pericyte1 may change the tissue biological properties, as ligament was dominant by fibroblast. To gain more biological insights underlying ECs, we performed Degs analysis between normal and degenerated groups (figure 4E). Combined with the result of enrichment analysis and GSEA analysis, we identified that ligamental ECs upregulated inflammation and complement related genes, such as C1QA, PLA2G2A, and ECM related genes such as PRG4, VIM, and POSTN during degeneration procession (figure 4H). The term “inflammation mediated by chemokine and cytokine signaling pathway” and “TGF-beta signaling pathway” activated in diseased group also implied that the inflammation and ECM remodeling have significant roles in the disease process (figure 4F).

**Figure 4:**
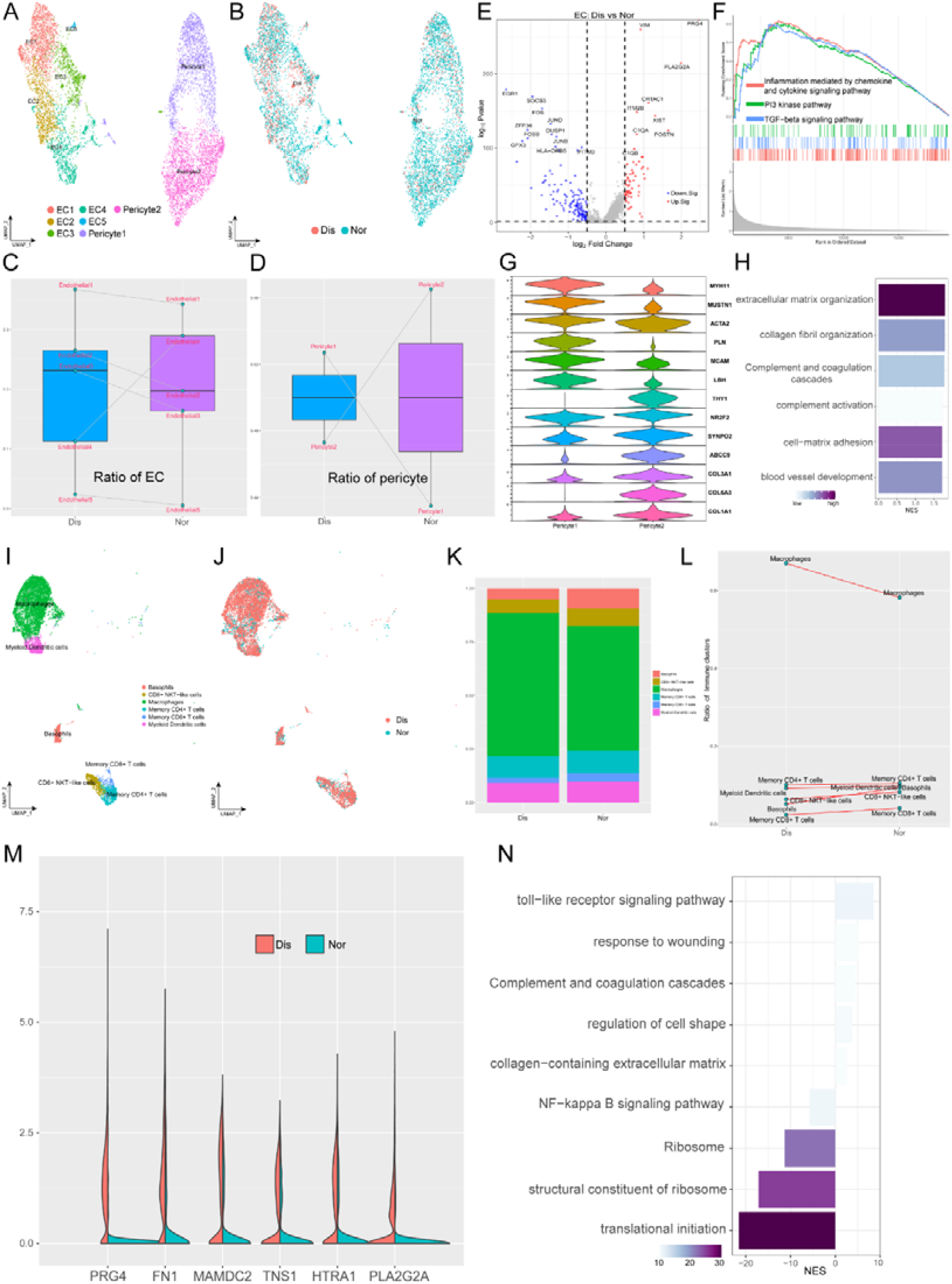
Identification of blood vessel derived cell and immune cell subclusters in human ligament. A: UMAP visualization of the subclusters of endothelial cell and pericyte. B: UMAP visualization of the distribution of endothelial cell and pericyte subclusters at different sample statuses. C and D: Summarized subpopulations of endothelial cell and pericyte percentage changes. E: Volcano plots displaying the DEGs in endothelial cell between normal group and diseased group. Each dot represented one gene. Red dots, differentially up-regulated genes; blue dots, differentially down-regulated genes; gray dot, non-differentially expressed genes. F: GSEA enrichment plots for representative signaling pathways upregulated in endothelial cell of diseased samples, compared with normal samples. G: Violin plots showing representative marker genes associated with different types of pericyte expressed in pericyte1 and pericyte2. H: GO and KEGG enrichment analysis of DEGs between normal and diseased endothelial cells. I: UMAP visualization of the subclusters of immune cell. J: UMAP visualization of the distribution of immune cell subclusters at different sample statuses. K: The proportion of each subcluster of immune cells in the lesioned and normal ligament. L: Summarized subpopulations of immune cell percentage changes. M: Violin plots showing representative genes of macrophages between normal and degenerated states. N: GO and KEGG enrichment analysis of DEGs between normal and diseased macrophages.

As an indispensable cellular component in the ligament, we identified six subpopulations of immune cells, including basophils, CD8^+^ NKT-like cells, macrophages, memory CD4^+^ T cells, memory CD8^+^ T cells, and myeloid dendritic cells (figure 4I and J). From figure 4K and L, we can conclude that macrophages are the most abundant immune cells in both normal and diseased groups and compared with the normal group, only the proportion of macrophages was increased in the degenerated group. We explored the gene profiling change of macrophages in the process of degeneration. As is shown in the violin plot figure 4M, we suggested that ECM remolding related genes have higher expression level in the diseased group, such as PRG4, FN1 and HTRA1. The enrichment analysis also implied that ECM organization and complement and coagulation cascades were activated in the degenerated ligament, which was basically consistent with the results of ECs analysis (figure 4N).

### 5. Putative signaling network for the intercellular crosstalk regulating the microenvironment homeostasis during ligamental degeneration

Elucidating the explicit interaction among fibroblasts, ECs and immune cells in the ligamental microenvironment will shed light on the mechanisms of the pathogenesis of ligamental degeneration. CellphoneDB and CellChat analysis were used to investigate the signaling network among the main cell clusters in the ligament. The heatmap illustrated the intensity of interactions among different cell clusters. From the results, we can find that in addition to interactions within fibroblast subclusters, cell-cell cross-talks were mainly between fibroblast subpopulations and ECs and between fibroblast subpopulations and macrophages. At the same time, we can find that fibroblast subsets dominated by the diseased group had stronger interaction associations with other cells, which implied that there are stronger and more complex interactions in degenerated ligaments (figure 5A).

**Figure 5:**
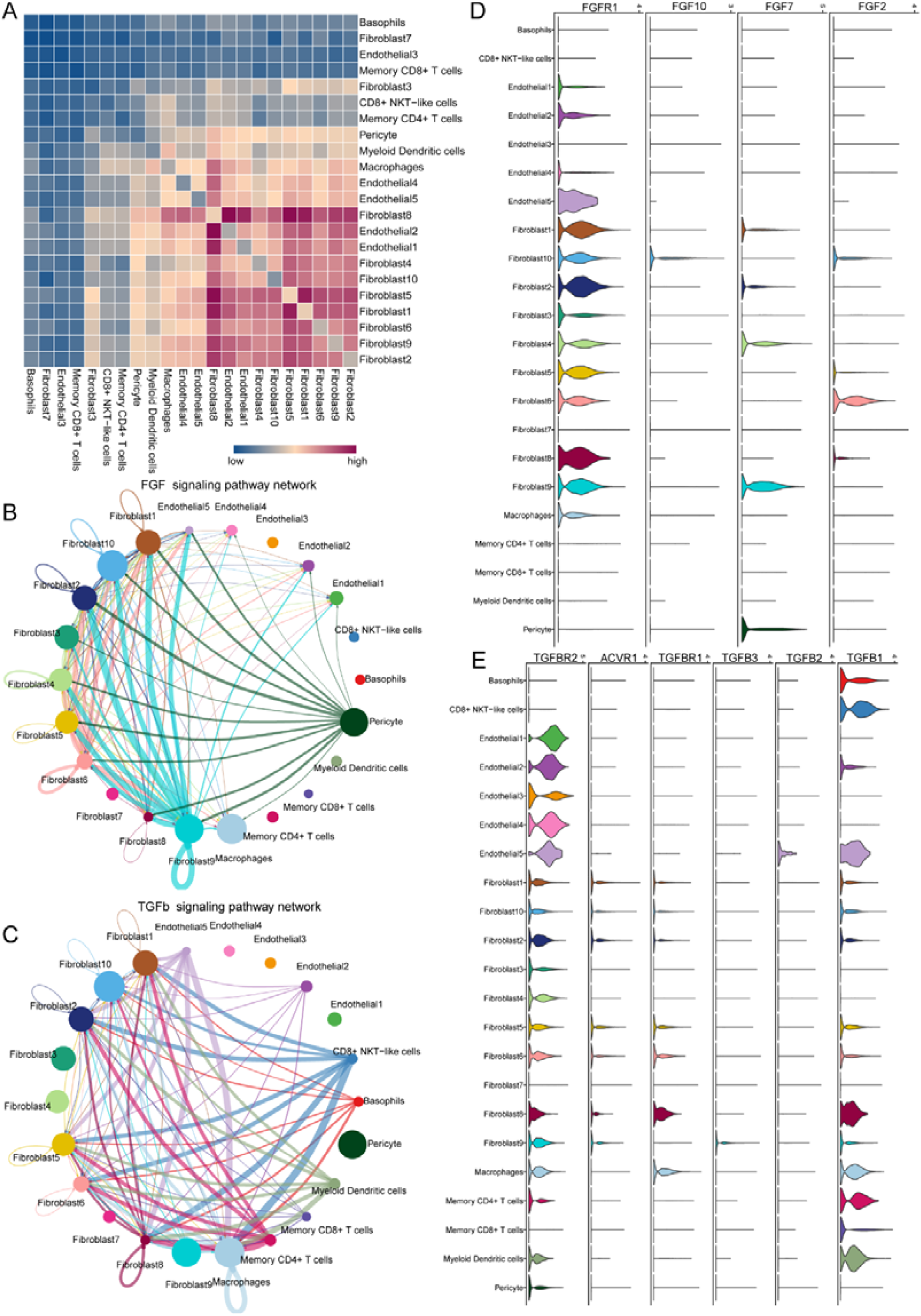
Cell–cell crosstalk during ligamental degeneration progression A: Heatmap depicting the significant interactions among the identified subclusters of fibroblast, immune cell, and endothelial cell. B and C: Circle plots showing the inferred TGF-β and FGF signaling networks. D and E: Violin plots showing ligand-receptor interactions related to TGF-β and FGF signaling pathways among subclusters of fibroblast, endothelial cell and immune cell.

To determine the important factors, we further analyzed the intercellular signaling networks of FGF and TGFB. Interestingly, Fib.9 was obviously involved in FGF signaling, both autocrine and paracrine (figure 5B). Fibroblasts were the leading receiver of FGF signals, as expected. Through the violin plot, we identified that Fib.9 acts on FGFR1 located on each fibroblast subpopulation by expressing FGF7 and FGF signaling pathway can promote pathological proliferation of cells and participate in scar repair (figure 5D). We inferred that the FGF signaling network may disrupt the homeostasis of fibroblasts in the ligaments and promote the progression of ligamental degeneration. Moreover, the TGF-β pathway was involved in many cell-cell interactions among fibroblasts subpopulations and macrophages via TGFB1-TGFBR1 or TGFB1-TGFBR2 (figure 5C and E). As shown above, fibroblasts of the degenerated group highly expressed PRG4, COL3A1, and SSP1 and these genes all important downstream target of TGF-β, which participate in the process of ECM remolding. Hence, we suggested that TGFB1 is a key gene and macrophages promote the development of the disease may through this molecule.

### 6. SpRNA-seq of ligament deciphering the spatial interactions of cell subclusters

For further insight, multi-angle interpretation of the cell composition changes that occurred in ligamental degeneration, we performed spRNA-seq on normal and lesioned ligaments. Firstly, by using feature marker transfer method, we mapped the individual subsets identified by single-cell sequencing into spatial transcriptome data. The heatmap illustrated that Fib.6, as a representative of fibroblasts in normal ligament tissue, was widely distributed in L1 and the amount was significantly higher than that in L8. The number of Fib.13, as a representative of fibroblasts in degenerated ligament, in L8 was obviously higher than that in L1 (figure 6A-D). We also examined the immune cells and ECs. As expected, there were more immune cells and ECs in L8 than in L1, implying immune infiltration and vascular hyperplasia were presented in the degenerative specimens (figure 6E-J). These results are consistent with those obtained by single cell sequencing. After that, we performed spotlight analysis in these two groups. All cell subsets are rendered in four colors, with red being the fibroblast subsets specific to the disease group. Finally, we hide the areas that are not red in both samples. We can find that the red region in L8 was widely distributed and the area was significantly higher than that in L1 group. And after careful observation, we found that almost all the red dots had yellow and black areas (figure 6K and L). This indicates that some fibroblast subpopulation specific to the diseased group are adjacent to endothelial and immune cells in the degenerative state. The proximity of different cells in space makes it possible for them to interact with each other. Therefore, this further provided strong evidence for the results of our single-cell interaction analysis

**Figure 6:**
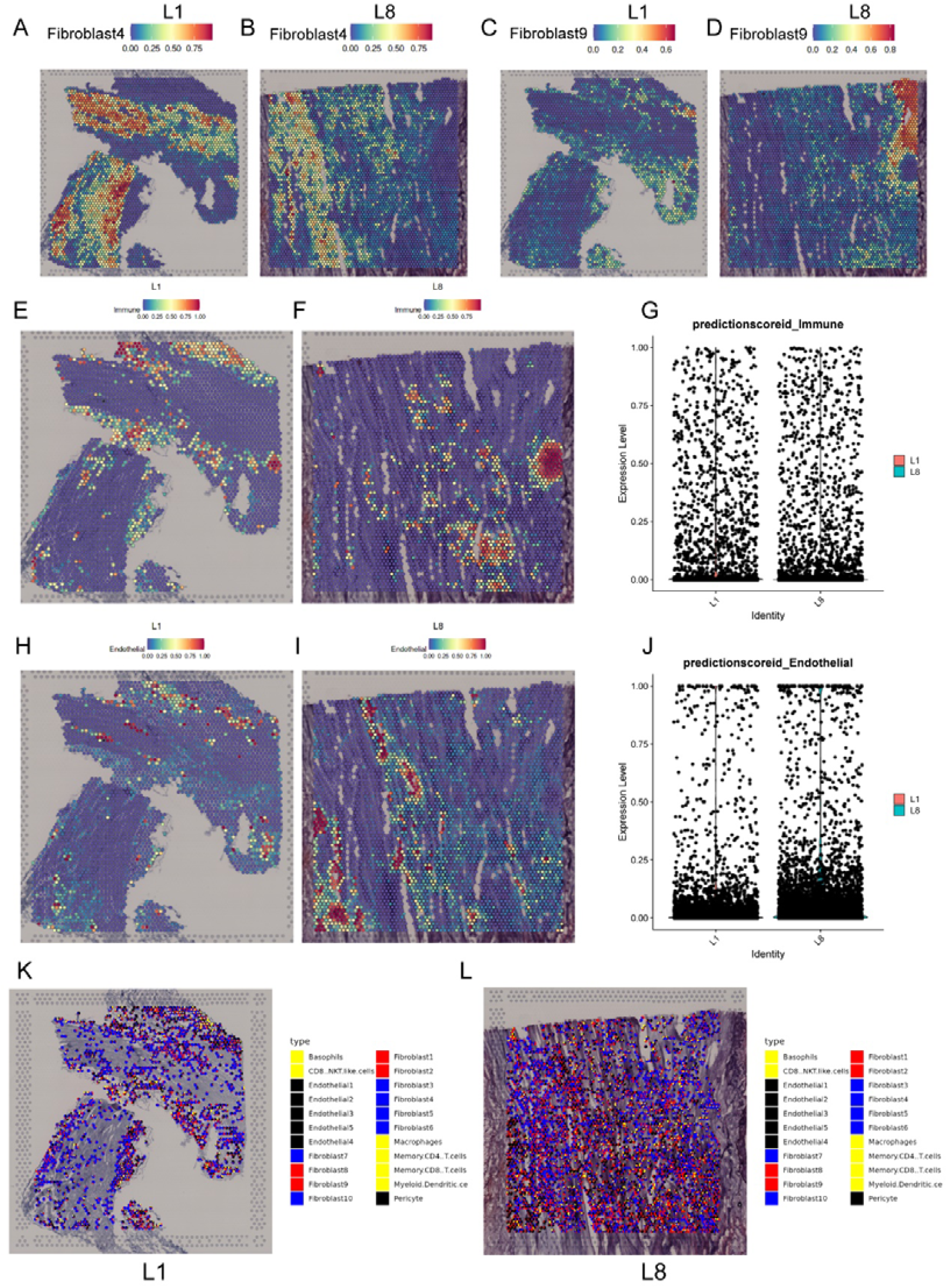
Spatial transcriptome sequencing deciphers the microenvironment changes during ligamental degeneration progression A-D: Spatial heatmaps showing the representative normal and degenerated fibroblast subcluster distribution in L1 and L8 samples. E and F: Spatial heatmaps showing the immune cells distribution in normal and degenerated ligamental samples. G: Quantitative analysis of immune cells in these two types of tissues. H and I: Spatial heatmaps showing the endothelial cells distribution in normal and degenerated ligamental samples. J: Quantitative analysis of endothelial cells in these two types of tissues. K and L: Spot light showing fibroblast subpopulation unique to diseased samples proportion in L1 and L8 samples.

## Discussion

ACL degeneration can contribute to cartilage injury and even OA onset and progression and may put a great burden on the normal life of many patients(Georgoulis et al., 2010; Hasegawa et al., 2012). But the mechanisms of this disease are not well characterized and treatments to prevent or treat ACL degeneration are scarce and not effective(Shane Anderson & Loeser, 2010; Tozer & Duprez, 2005). Healthy and degenerated ACL tissues include multiple cell subpopulations with diverse genetic and phenotypic characteristics. How this heterogeneity emerges in development degeneration remains unclear. Herein, we built a single-cell atlas of normal and degenerated human ACL and explored the characteristics and key regulatory pathways of distinct fibroblast subtypes. These findings will help us understand the pathogenesis of ACL degeneration in depth, and provide potential targets for clinical therapies of this disease.

Fibroblasts are increasingly considered as dominating cell types in ligaments and central mediators of ligamental degeneration, and here we identified 10 fibroblast subpopulations in human normal and degenerated ACL samples by using scRNA-seq. Further cluster analysis profiled characteristics of each subclusters and combined with the cell proportion analysis we found that pro-inflammation and ECM remodeling related fibroblast subgroups were significantly increased in the degenerative ligamental tissues, which implied that inflammation and ECM remodeling are key events in the disease process. Through comparing the Degs of fibroblasts between degenerative and healthy conditions, we found that ECM related genes were upregulated in the diseased group such as FN1, COL3A1, PRG4 and SPP1. The results of enrichment analysis and GSEA also illustrated that ECM related pathways were activated in the degenerated group, which suggested that the modules of ECM play an important role in the process of ligamental degeneration. According to the gene interaction analysis, two gene modules were activated in the diseased group, leukocyte trans-endothelial migration and ECM-receptor interaction. These findings were consistent with previous studies which suggested that ligament inflammation and ECM changes contribute to ACL degeneration(Busch et al., 2013; Hasegawa et al., 2012). It is due to the changes and remodeling of the ECM in the degenerative ligament tissue that the biomechanical changes of the ligament are caused and eventually may result in the rupture of the ligament. From the pseudotime analysis, we identified two cell fates in the ligamental degeneration. Cell fate 1 represents a progressive process of disease characterized by inflammatory damage and extracellular matrix degradation. Cell fate 2 implies a chronic repair process for the disease characterized by ECM remodeling, ligament development, and damage repair.

Immune cells are closely related to ligamental degeneration(Kim-Wang et al., 2021). We analyzed the heterogeneity of immune cells in ACL using single-cell RNA sequencing. The results illustrated that macrophages are dominating immune cells in ACL and inflammation related genes PLA2G2A and ECM related genes PRG4, FN1, and HTRA1 were upregulated in the macrophages of the diseased group. Enrichment analysis also revealed that complement and ECM related pathways were activated in the degenerated group. This implied that macrophages highly expressed ECM and inflammation associated genes in the ligamental degeneration progression. So, we suggested that macrophages may contribute to the ligamental degeneration. We also identified two types of pericytes in these tissues. As we all know, pericyte in the tissue have some stem cell characteristics and can differentiate into fibroblasts, and chondrocytes(Armulik, Genové, & Betsholtz, 2011; Smyth et al., 2018). Pericyte 1 highly expressed stem cell related genes MCAM, MUSTN1 and have the characteristics of myofibroblasts and pericyte 2 tend to have the characteristics of fibroblasts. Pericyte 1 was increased and pericyte 2 decreased in the degenerated group, which may change the properties of ligamental cells and contribute to the ligamental degeneration.

Cells can communicate via ligand-receptor interactions(Kumar et al., 2018), so targeting cell-cell interactions is frequently utilized in the clinical treatment. The results of CellphoneDB analysis demonstrated that the high content of fibroblasts and immune cells in the degenerative group had significantly higher interaction intensity, such as fibroblast 1,2, and 8, which means that the interaction between cells in the ligament was enhanced in the degenerative state. To identify the key factors regulating the disease process, CellChat analysis was used to dissect the intercellular crosstalk based on the signaling network in the human ACL. We found that FGF signaling pathway and TGF-β signaling pathway were involved in the crosstalk network. TGFβ1, contributes to osteophyte formation by inducing endochondral ossification, is thought to be closely related to the onset of OA(Murata et al., 2019) and can be used to induce chondrogenic differentiation of ligament-derived stem cells in vitro(Schwarz et al., 2019). It has been reported that in myocardial fibrosis, liver fibrosis, glaucoma and other diseases, TGF-β signaling pathway is also involved in angiogenesis, ECM remodeling, pathological scar healing(Ilieş et al., 2021; Lee & Massagué, 2022; P et al., 2018; Penn, Grobbelaar, & Rolfe, 2012). FGFs can induce the fibroblast to myofibroblast differentiation, promote tissue repair by regulating cell proliferation, survival and angiogenesis and contribute to ECM remodeling(Kendall & Feghali-Bostwick, 2014; Ma, Iyer, Jung, Czubryt, & Lindsey, 2017). ECM remodeling by overexpression, degradation and cross-linking of ECM proteins in the lesioned tissue are direct factors leading to fibrosis tissues(Sun et al., 2022). In this study, we observed that fibroblast9, inflammation related fibroblast, highly expressed FGF7 and can act on FGFR1 mainly distributed in the fibroblast subgroups. Immune cells especially macrophages highly expressed TGFB1 and can act on TGFBR1 and TGFBR2 mainly distributed in the fibroblast and endothelial subgroups. These findings suggested that FGF and TGF-β signaling pathways may induce ECM remolding in the process of ligamental degeneration and FGF7-FGFR1 and TGFB1-TGFBR2 may be potential targets for the treatment of this disease.

SpRNA-seq was used to detect the spatial information underlying the normal and degenerated ligaments and validate the findings acquiring from the ScRNA-seq. Through the results, we observed that the disease group specific fibroblasts and immune cells recognized by single cells were more numerous and more widely distributed in the disease group specimens. We also get some spatial information, that fibroblasts in the disease group were spatially closer to immune and endothelial cells, which was more conducive to cell interaction.

## Conclusions

In conclusion, our study described the cell atlas of the human ligament, providing a valuable resource for further investigate of ACL homeostasis and the pathogenesis of ligamental degeneration. The cellular heterogeneity and signaling network we uncovered help to increase the understanding of the human ACL at a single-cell level and provide crucial clues for establishing new diagnostic and therapeutic strategies for this disease in the future.

## Data Availability

Data are available in a public, open-access repository. The single-cell RNA-seq data and cluster annotations are available at GSA for human (https://ngdc.cncb.ac.cn/gsa-human/) with the accession number PRJCA014157.

## Declarations

### Ethics approval and consent to participate

This study was reviewed and approved by our University Ethics Committee (Ethics Committee on Biomedical Research, West China Hospital of Sichuan University No. 658 2020-(921)) and all procedures complied with the Helsinki Declaration. Participants gave informed consent to participate in the study.

### Consent for publication

Not applicable.

### Author’s Contributions

WF and JL conceived the project. WF and JL designed the experiments. WF, RY and MG collected the specimens. RY, TX, LZ and LY performed bioinformatic and statistical analyses. WF and RY wrote the manuscripts. All authors commented and revised the manuscripts.

## Acknowledgements

We appreciate all patients who participated in this study.

## Figure legends

**Supplementary figure 1:**
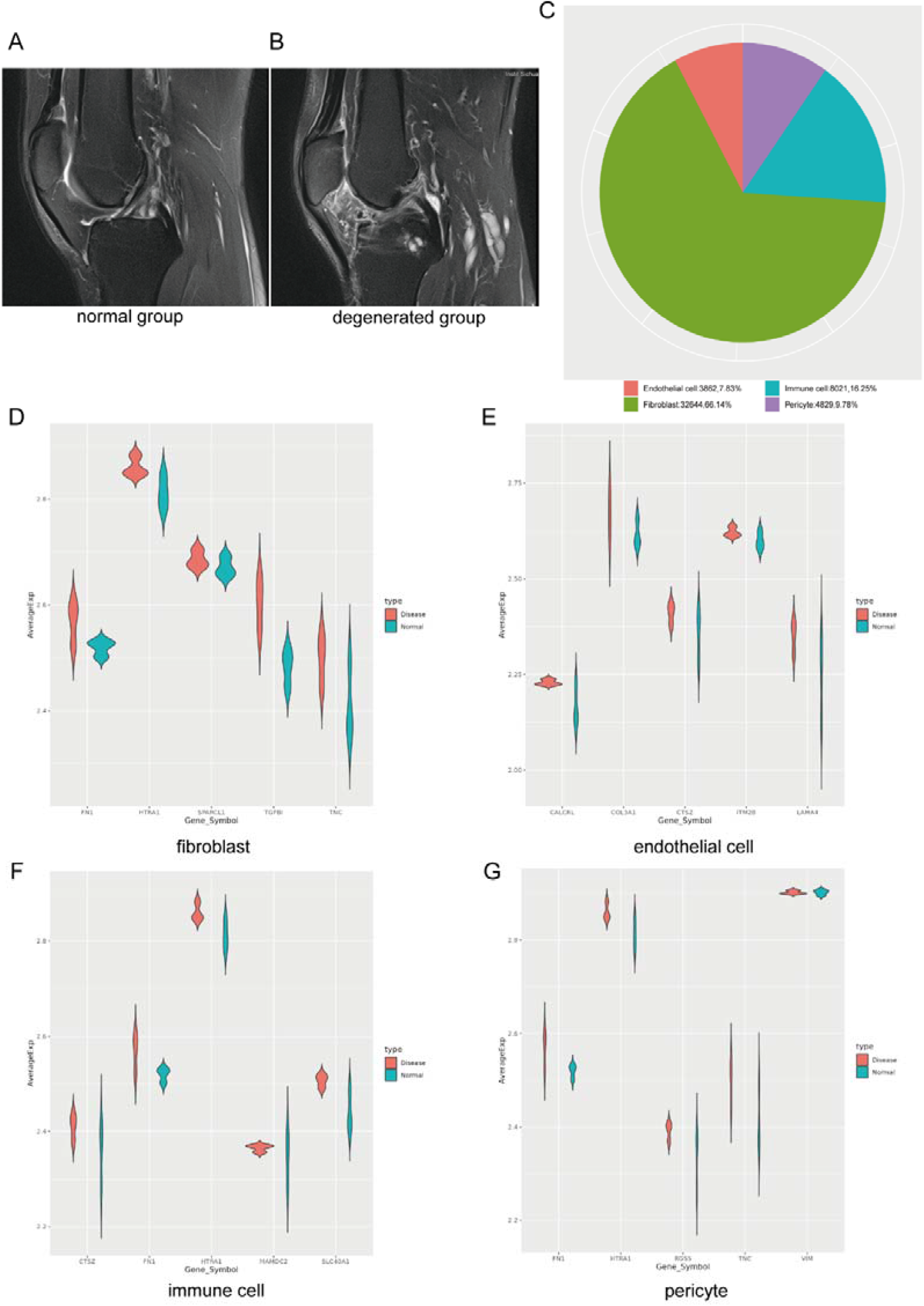
A and B: MRI Photographs of typical normal and degenerated ACLs. C: The proportion of each cell type. D-G: The expression of the top five genes highly expressed in degenerated state obtained by single cell sequencing of fibroblast, endothelial cell, immune cell and pericyte in bulk sequencing data.

## References

Armulik, A., Genové, G., & Betsholtz, C. (2011). Pericytes: developmental, physiological, and pathological perspectives, problems, and promises. Dev Cell, 21(2), 193–215. doi:10.1016/j.devcel.2011.07.001

Beard, J. R., & Bloom, D. E. (2015). Towards a comprehensive public health response to population ageing. Lancet, 385(9968), 658–661. doi:10.1016/s0140-6736(14)61461-6

Briggs, A. M., Cross, M. J., Hoy, D. G., Sànchez-Riera, L., Blyth, F. M., Woolf, A. D., & March, L. (2016). Musculoskeletal Health Conditions Represent a Global Threat to Healthy Aging: A Report for the 2015 World Health Organization World Report on Ageing and Health. Gerontologist, 56 Suppl 2, S243–255. doi:10.1093/geront/gnw002

Busch, C., Girke, G., Kohl, B., Stoll, C., Lemke, M., Krasnici, S., … Schulze-Tanzil, G. (2013). Complement gene expression is regulated by pro-inflammatory cytokines and the anaphylatoxin C3a in human tenocytes. Mol Immunol, 53(4), 363–373. doi:10.1016/j.molimm.2012.09.001

Cao, J., Spielmann, M., Qiu, X., Huang, X., Ibrahim, D. M., Hill, A. J., … Shendure, J. (2019). The single-cell transcriptional landscape of mammalian organogenesis. Nature, 566(7745), 496–502. doi:10.1038/s41586-019-0969-x

Corps, A. N., Robinson, A. H., Movin, T., Costa, M. L., Hazleman, B. L., & Riley, G. P. (2006). Increased expression of aggrecan and biglycan mRNA in Achilles tendinopathy. Rheumatology (Oxford), 45(3), 291–294. doi:10.1093/rheumatology/kei152

Fleming, B. C. (2003). Biomechanics of the anterior cruciate ligament. J Orthop Sports Phys Ther, 33(8), A13–15.

Frank, C. B. (2004). Ligament structure, physiology and function. J Musculoskelet Neuronal Interact, 4(2), 199–201.

Georgoulis, A. D., Ristanis, S., Moraiti, C. O., Paschos, N., Zampeli, F., Xergia, S., … Mitsionis, G. (2010). ACL injury and reconstruction: Clinical related in vivo biomechanics. Orthop Traumatol Surg Res, 96(8 Suppl), S119–128. doi:10.1016/j.otsr.2010.09.004

Hasegawa, A., Nakahara, H., Kinoshita, M., Asahara, H., Koziol, J., & Lotz, M. K. (2013). Cellular and extracellular matrix changes in anterior cruciate ligaments during human knee aging and osteoarthritis. Arthritis Res Ther, 15(1), R29. doi:10.1186/ar4165

Hasegawa, A., Otsuki, S., Pauli, C., Miyaki, S., Patil, S., Steklov, N., … Lotz, M. K. (2012). Anterior cruciate ligament changes in the human knee joint in aging and osteoarthritis. Arthritis Rheum, 64(3), 696–704. doi:10.1002/art.33417

Hayashi, K., Frank, J. D., Hao, Z., Schamberger, G. M., Markel, M. D., Manley, P. A., & Muir, P. (2003). Evaluation of ligament fibroblast viability in ruptured cranial cruciate ligament of dogs. Am J Vet Res, 64(8), 1010–1016. doi:10.2460/ajvr.2003.64.1010

Ilie□, R. F., Aioanei, C. S., Catana, A., Halmagyi, S. R., Lukacs, I., Tokes, R. E., … Pop, I. V. (2021). Involvement of COL5A2 and TGF-β1 in pathological scarring. Exp Ther Med, 22(4), 1067. doi:10.3892/etm.2021.10501

Kendall, R. T., & Feghali-Bostwick, C. A. (2014). Fibroblasts in fibrosis: novel roles and mediators. Front Pharmacol, 5, 123. doi:10.3389/fphar.2014.00123

Kharaz, Y. A., Canty-Laird, E. G., Tew, S. R., & Comerford, E. J. (2018). Variations in internal structure, composition and protein distribution between intra- and extra-articular knee ligaments and tendons. J Anat, 232(6), 943–955. doi:10.1111/joa.12802

Kim-Wang, S. Y., Holt, A. G., McGowan, A. M., Danyluk, S. T., Goode, A. P., Lau, B. C., … McNulty, A. L. (2021). Immune cell profiles in synovial fluid after anterior cruciate ligament and meniscus injuries. Arthritis Res Ther, 23(1), 280. doi:10.1186/s13075-021-02661-1

Kumar, M. P., Du, J., Lagoudas, G., Jiao, Y., Sawyer, A., Drummond, D. C., … Raue, A. (2018). Analysis of Single-Cell RNA-Seq Identifies Cell-Cell Communication Associated with Tumor Characteristics. Cell Rep, 25(6), 1458-1468.e1454. doi:10.1016/j.celrep.2018.10.047

Laurencin, C. T., & Freeman, J. W. (2005). Ligament tissue engineering: an evolutionary materials science approach. Biomaterials, 26(36), 7530–7536. doi:10.1016/j.biomaterials.2005.05.073

Lee, J. H., & Massagué, J. (2022). TGF-β in Developmental and Fibrogenic EMTs. Semin Cancer Biol. doi:10.1016/j.semcancer.2022.09.004

Li, X., & Wang, C. Y. (2021). From bulk, single-cell to spatial RNA sequencing. Int J Oral Sci, 13(1), 36. doi:10.1038/s41368-021-00146-0

Loeser, R. F. (2010). Age-related changes in the musculoskeletal system and the development of osteoarthritis. Clin Geriatr Med, 26(3), 371–386. doi:10.1016/j.cger.2010.03.002

Ma, Y., Iyer, R. P., Jung, M., Czubryt, M. P., & Lindsey, M. L. (2017). Cardiac Fibroblast Activation Post-Myocardial Infarction: Current Knowledge Gaps. Trends Pharmacol Sci, 38(5), 448–458. doi:10.1016/j.tips.2017.03.001

Murata, K., Kokubun, T., Onitsuka, K., Oka, Y., Kano, T., Morishita, Y., … Kanemura, N. (2019). Controlling joint instability after anterior cruciate ligament transection inhibits transforming growth factor-beta-mediated osteophyte formation. Osteoarthritis Cartilage, 27(8), 1185–1196. doi:10.1016/j.joca.2019.03.008

P, M. F., L, M. N., O, C. M., M, G. L., Pereira, M. C., de Mendonça-Lima, L., … L, R. G. (2018). Inhibition of TGF-β pathway reverts extracellular matrix remodeling in T. cruzi-infected cardiac spheroids. Exp Cell Res, 362(2), 260–267. doi:10.1016/j.yexcr.2017.11.026

Penn, J. W., Grobbelaar, A. O., & Rolfe, K. J. (2012). The role of the TGF-β family in wound healing, burns and scarring: a review. Int J Burns Trauma, 2(1), 18–28.

Roos, H., Adalberth, T., Dahlberg, L., & Lohmander, L. S. (1995). Osteoarthritis of the knee after injury to the anterior cruciate ligament or meniscus: the influence of time and age. Osteoarthritis Cartilage, 3(4), 261–267. doi:10.1016/s1063-4584(05)80017-2

Schulze-Tanzil, G. (2019). Intraarticular Ligament Degeneration Is Interrelated with Cartilage and Bone Destruction in Osteoarthritis. Cells, 8(9). doi:10.3390/cells8090990

Schwarz, S., Gögele, C., Ondruschka, B., Hammer, N., Kohl, B., & Schulze-Tanzil, G. (2019). Migrating Myofibroblastic Iliotibial Band-Derived Fibroblasts Represent a Promising Cell Source for Ligament Reconstruction. Int J Mol Sci, 20(8). doi:10.3390/ijms20081972

Shane Anderson, A., & Loeser, R. F. (2010). Why is osteoarthritis an age-related disease? Best Pract Res Clin Rheumatol, 24(1), 15–26. doi:10.1016/j.berh.2009.08.006

Smyth, L. C. D., Rustenhoven, J., Scotter, E. L., Schweder, P., Faull, R. L. M., Park, T. I. H., & Dragunow, M. (2018). Markers for human brain pericytes and smooth muscle cells. J Chem Neuroanat, 92, 48–60. doi:10.1016/j.jchemneu.2018.06.001

Sun, C., Tian, X., Jia, Y., Yang, M., Li, Y., & Fernig, D. G. (2022). Functions of exogenous FGF signals in regulation of fibroblast to myofibroblast differentiation and extracellular matrix protein expression. Open Biol, 12(9), 210356. doi:10.1098/rsob.210356

Thompson, S., Salmon, L., Waller, A., Linklater, J., Roe, J., & Pinczewski, L. (2015). Twenty-year outcomes of a longitudinal prospective evaluation of isolated endoscopic anterior cruciate ligament reconstruction with patellar tendon autografts. Am J Sports Med, 43(9), 2164–2174. doi:10.1177/0363546515591263

Tozer, S., & Duprez, D. (2005). Tendon and ligament: development, repair and disease. Birth Defects Res C Embryo Today, 75(3), 226–236. doi:10.1002/bdrc.20049

Wen, L., & Tang, F. (2018). Boosting the power of single-cell analysis. Nat Biotechnol, 36(5), 408–409. doi:10.1038/nbt.4131

Zeng, Y., He, J., Bai, Z., Li, Z., Gong, Y., Liu, C., … Liu, B. (2019). Tracing the first hematopoietic stem cell generation in human embryo by single-cell RNA sequencing. Cell Res, 29(11), 881–894. doi:10.1038/s41422-019-0228-6

